# High Ambient Temperature in Pregnancy and Risk of Childhood Acute Lymphoblastic Leukemia

**DOI:** 10.1101/2023.05.19.23290227

**Authors:** Tormod Rogne, Rong Wang, Pin Wang, Nicole C. Deziel, Catherine Metayer, Joseph L. Wiemels, Kai Chen, Joshua L. Warren, Xiaomei Ma

## Abstract

**Background:** High ambient temperature is increasingly common due to climate change and is associated with risk of adverse pregnancy outcomes. Acute lymphoblastic leukemia (ALL) is the most common malignancy in children, the incidence is increasing, and in the United States it disproportionately affects Latino children. We aimed to investigate the potential association between high ambient temperature in pregnancy and risk of childhood ALL.

**Methods:** We used data from California birth records (1982-2015) and California Cancer Registry (1988-2015) to identify ALL cases diagnosed <14 years and 50 times as many controls matched by sex, race/ethnicity, and date of last menstrual period. Ambient temperatures were estimated on a 1-km grid. Association between ambient temperature and ALL was evaluated per gestational week, restricted to May-September, adjusting for confounders. Bayesian meta-regression was applied to identify critical exposure windows. For sensitivity analyses, we evaluated a 90-day pre-pregnancy period (assuming no direct effect before pregnancy) and constructed an alternatively matched dataset for exposure contrast by seasonality.

**Findings:** Our study included 6,258 ALL cases and 307,579 controls. The peak association between ambient temperature and risk of ALL was observed in gestational week 8, where a 5 °C increase was associated with an odds ratio of 1.09 (95% confidence interval 1.04-1.14) and 1.05 (95% confidence interval 1.00-1.11) among Latino and non-Latino White children, respectively. The sensitivity analyses supported this.

**Interpretation:** Our findings suggest an association between high ambient temperature in early pregnancy and risk of childhood ALL. Further replication and investigation of mechanistic pathways may inform mitigation strategies.

## INTRODUCTION

Climate change in the form of high ambient temperature has had devastating consequences for human health worldwide and is projected to have an even greater impact in the future.^1^ Racial/ethnic minority groups experience a disproportionate burden of heat exposure, in part due to line of work and residential segregation.^1–3^ It is increasingly clear that high ambient temperature during pregnancy has negative effects on birth outcomes, especially in racial/ethnic minority groups.^4,5^ However, very little is known about the longer-term outcomes for the offspring as a consequence of exposure to high ambient temperature during pregnancy. ALL is the most common childhood malignancy with more than 50,000 new cases every year worldwide, and the incidence is steadily increasing.^6^ In the United States (US), there is considerable disparity by race/ethnicity, where Latino children have a roughly 40% higher risk of ALL compared with non-Latino White children, and Latino children also have the greatest yearly increase in ALL incidence of any racial/ethnic group.^6,7^ The large difference in ALL risk observed between Latino and non-Latino children is not explained by genetic differences.^8^

It is well established that most cases of childhood ALL have a prenatal origin,^9,10^ and environmental exposures occurring during pregnancy such as air pollution have been associated with an increased risk of childhood ALL.^11–13^ Exposure during the first trimester is suspected to be the most critical because this is when the most profound developmental alterations in hematopoiesis occur (lymphopoiesis starts around gestational week 8).^14^ There are multiple reasons to suspect that maternal exposure to high ambient temperature may initiate the pathogenesis of ALL in fetal life. First, pre-leukemic clones may be caused by oxidative stress,^9^ and high ambient temperature increases oxidative stress.^15^ Also, inflammatory markers, heat shock proteins, and stress hormones increase in response to heat,^16^ and stress in pregnancy has been linked to an increased risk of ALL and other childhood cancers.^17,18^

Using a population-based linkage study derived from the general birth cohort in California (1982-2015), we conducted a nested case-control study to evaluate the potential association between high ambient temperature during pregnancy and risk of ALL in the offspring. In particular, we aimed to identify critical windows of exposure and to evaluate possible differences by racial/ethnic groups. We hypothesized that any harmful effect of high ambient temperature would be the greatest during the first trimester and that Latino children would be impacted more by high temperatures. This is the first study to directly evaluate the association between ambient temperature in pregnancy and the risk of cancer in the offspring.

## METHODS

### Study Population

This study leveraged the California Linkage Study of Early-Onset Cancers (CALSEC), which is a population-based, statewide linkage study that included children born in California during 1982-2015 and diagnosed with childhood cancer in California during 1988-2015.^12,19^ Non-cancer controls were selected from the statewide birth records. Information on cancer status was gathered from the California Cancer Registry, and birth records from the Center for Health Statistics and Informatics, both within the California Department of Public Health. This study received approval from the institutional review boards at California Department of Public Health, Yale University, University of California, Berkeley, and University of Southern California.

We used the International Classification of Diseases for Oncology, 3^rd^ edition, to identify cases, where codes of 9811-9818, 9826, 9835-9837 were considered consistent with a diagnosis of ALL. An age-limit of <14 years was used to define childhood ALL.^7^ We initially identified 6,849 cases of childhood ALL. Of these, we excluded children who 1) had missing residential address at the time of birth (n=142); 2) had mothers who resided outside of California (n=7); 3) had missing information on gestational age (n=349), birth weight (n=1), birth order (n=3), mother’s birthplace (n=4), or delivery mode (n=3); or 4) had congenital abnormalities (n=79) or had missing information on this item (n=2). We next selected 50 (or as many available meeting our criteria) non-cancer controls per case matched on sex, race/ethnicity, and date of mother’s last menstrual period (LMP) ±7 days. One case was further excluded due to no available matched controls. A total of 6,258 cases and 307,579 controls were included in the primary analyses of this study.

### Temperature Ascertainment

Maternal residential addresses at the time of her child’s birth were geocoded.^19^ Exposure to ambient temperature was estimated from the Daily Surface Weather Data on a 1-km Grid for North America, Version 4.^20^ This dataset utilizes a combination of interpolation and extrapolation and multiple inputs from weather stations and provides information on daily minimum and maximum temperature at a spatial resolution of 1 km. The exposure of interest was daily mean temperature which was estimated from daily minimum and maximum temperatures. The smallest unit of exposure used in our analyses was the mean weekly ambient temperature, which was estimated based on the average daily mean temperature each week.

### Other Variables

Birth records were used to gather the following information about our study population: Gestational age at birth (in days), date of birth, race/ethnicity (Latino, non-Latino White, non-Latino Black, non-Latino Asian/Pacific Islander, other), sex, birth order (1^st^, 2^nd^, >=3^rd^), maternal age (<20, 20-24, 25-29, 30-34, >=35 years old), paternal age (<25, 25-29, 30-34, 35-39, >=40 years old), maternal education (<8, 9-11, 12, 13-15, >=16 years), birth weight (grams), and mode of delivery (vaginal or cesarean section). Maternal residential addresses were linked to 2000 Census block group data to obtain the proportion in the block group living below the 150% federal poverty level (tertiles based on the distribution among controls). Date of the first day of LMP was estimated based on date of birth and registered gestational age at birth.

### Statistical Analyses

This study included a range of analyses that are described in detail below. In brief, the main analysis evaluated the gestational week-specific association between high ambient temperature and risk of ALL after adjusting for key confounders and matching on race/ethnicity, sex and date of LMP. We also evaluated subgroups and conducted non-linear analyses. Finally, we conducted two important sensitivity analyses: 1) The association between high ambient temperature *before* pregnancy and risk of ALL (i.e., a form of negative control exposure); and 2) analyses on an alternatively matched dataset matching on *residential address* and *year* of LMP instead of *date* of LMP (i.e., to evaluate exposure contrast by seasonality instead of location).

We evaluated the association between weekly mean ambient temperature and the risk of childhood ALL, one week at a time before running Bayesian meta-regression analyses (see below). All results are expressed as odds ratio (OR) for ALL per 5 °C increase in mean weekly ambient temperature. Given that we wanted to evaluate the effect of high ambient temperature, we restricted the season of interest to the warm period May 1^st^-September 30^th^. For any given week of exposure, at least one day of that week had to be in the warm period.

One of the objectives of our study was to identify whether there were critical windows of exposure. Thus, we considered ambient exposure to temperature in each week of pregnancy from LMP to date of birth in separate models. Additionally, we evaluated the 13 weeks (90 days) preceding LMP as an approximate negative control exposure period.^21^ As embryonic lymphopoiesis starts around gestational week 8,^14^ we hypothesized that we would see no association between ambient temperature and ALL in the weeks preceding pregnancy.

In our analyses, we adjusted for the matching variables (i.e., race/ethnicity, sex and date of LMP ±7 days) and a set of additional potential confounders identified through literature search and construction of directed acyclic graphs: Birth order, maternal and paternal age, maternal education, neighborhood poverty, seasonality and time trend (captured through LMP), and residential address at the time of birth. Given that exposure contrast to ambient temperature is achieved by time and/or space, we could not simultaneously match on date of LMP *and* residential address. Instead, we considered date of LMP to be the more important confounder of the two, which was therefore included in the primary analyses, while adjustment for residential address was achieved in a sensitivity analysis of a secondary matched dataset (see below). Unless stated otherwise, all characteristics and analyses presented were based on the primary matched dataset (i.e., matched on sex, race/ethnicity and date of mother’s LMP ±7 days).

We considered pregnancy complications and birth outcomes to be potential mediators of the association between high ambient temperature in pregnancy and risk of ALL. Similarly, air pollution is generally considered a mediator of the association between high ambient temperature and adverse health outcomes.^15^ We did not adjust for mediators.

### Evaluation of Critical Windows of Exposure

To identify critical windows of exposure, we analyzed our data in two stages. First, we ran conditional logistic regression models, separately for each week, where the association between mean ambient temperature and ALL risk was estimated, along with a standard error for the estimate. Next, we used a Bayesian meta-regression framework, where the estimates and standard errors from the first stage were used as input for the second analysis stage.

For the second analysis stage, we extended the original critical window variable selection (CWVS) method of Warren *et al*. to the meta-regression setting.^22^ The first stage association estimates were assumed to be normally distributed with mean equal to the true but unobserved association of interest, and variance equal to the squared standard error. The true associations were then modeled using the original CWVS methodology, which decomposes the single effect into continuous and binary components and uses a joint Gaussian process with temporal correlation structure to provide “smoothed” parameter estimation while conducting Bayesian variable selection output (Supplementary Methods). From this Bayesian analysis, we calculated posterior means as the point estimates and 95% quantile-based credible intervals to quantify uncertainty in the parameters (i.e., comparable to confidence intervals in the frequentist analysis setting). For consistency with the other analyses, the credible intervals are reported as confidence intervals.

### Subgroup Analyses

After identifying the gestational week where the association between ambient temperature and risk of ALL was the greatest (i.e., the largest estimated association that was statistically significant), we used this gestational week as basis for two subsequent subgroup analyses. First, we conducted analyses separately for Latino and non-Latino White subjects; the other racial/ethnic groups had too few cases to warrant stratified analyses. The other subgroup analysis was stratified on the age of diagnosis. Subgroup differences were compared by calculating the z-statistic.^23^

### Non-Linear Analysis

We conducted a regression analysis that included mean ambient temperature modeled using a restricted cubic spline with 5 knots (5^th^, 27.5^th^, 50^th^, 72.5^th^ and 95^th^ percentiles). This analysis was adjusted for the same confounders as the main analysis. The non-linear analysis was only carried out in the gestational week with the largest estimated association with ambient temperature exposure that was statistically significant. A log-likelihood ratio test was run to evaluate whether the non-linear model was a better fit than the linear model.

### Secondary Matched Dataset

To address potential residual confounding due to residential address at birth, we created a secondary matched dataset where we matched cases and controls on *year of LMP*, sex, race/ethnicity, and *residential address at birth within 10 km*. Thus, the difference between the main dataset and the secondary matched dataset was that the former compared pregnancies occurring at the same time (±7 days) but at different locations, while the secondary matched dataset compared pregnancies occurring in the same location (within 10 km) but at different times during the same year (Supplementary Figure 1). This analysis was not restricted to the warm period, but was otherwise adjusted for the same confounders as in the other analyses. Due to the strict matching criteria, the secondary matched dataset included four controls per case, and not all cases had available controls. In total, 6,188 cases and 24,434 controls were included in the secondary matched dataset.

Our hypothesis was that the secondary matched dataset would reveal a similar positive association between ambient temperature in the first trimester and risk of ALL. Given that this dataset was not restricted to the warm season, and as it was matched on year and location but not date of LMP, we expected to see an artificial inverse association roughly 6 months after the peak positive association (i.e., in the third trimester): An increased proportion of cases (compared with controls) conceived in early summer would result in higher mean ambient temperature (compared with controls) in early gestational weeks, while the ambient temperature would be lower among cases (compared with controls) in gestational weeks in late pregnancy as these would to a greater extent (compared with controls) be in the winter months.

### Software

The two-stage meta-regression analyses used the rjags package (version 4.13) in R (version 4.2.2), and all other analyses were performed with SAS version 9.4 (SAS Institute Inc., Cary, NC), with a statistical significance level of 0.05.

## RESULTS

For the warm season in the years 1982-2015, the mean of the weekly mean ambient temperature was 21.5 °C (standard deviation 3.7 °C), the 25^th^ and 75^th^ percentiles were 19.0 °C and 23.9 °C, respectively, and the minimum and maximum weekly mean temperatures were 0.0 °C and 40.1 °C, respectively.

A majority of the study population were male, and most were of Latino ethnicity (Table 1). Sex, race/ethnicity and year of birth were equally distributed between cases and controls as per matching design. Compared with controls, cases had slightly greater birth weight and were more likely to have mothers with higher education. Among the 6,258 cases, 3,636 (58.1%) were diagnosed before 5 years of age, 1,769 (28.3%) at the age of 5-9 years, and 853 (13.6%) between 10 and 14 years of age.

**Table 1.** Characteristics of Study Population

|  | Entire study population | Cases | Controls |
| --- | --- | --- | --- |
| **Female sex, n (%)** | 139,144 (44.3) | 2,773 (44.3) | 136,371 (44.3) |
| **Race/ethnicity, n (%)** |  |  |  |
| Latino | 174,906 (55.7) | 3,431 (54.8) | 171,475 (55.7) |
| Non-Latino White | 96,562 (30.8) | 1,898 (30.3) | 94,664 (30.8) |
| Non-Latino Black | 10,121 (3.2) | 223 (3.6) | 9,898 (3.2) |
| Non-Latino Asian/Pacific Islander | 30,959 (9.9) | 622 (9.9) | 30,337 (9.9) |
| Other | 1,289 (0.4) | 84 (1.3) | 1,205 (0.4) |
| **Year of birth, n (%)** |  |  |  |
| 1982-1995 | 135,097 (43.0) | 2,661 (42.5) | 132,436 (43.1) |
| 1996-2005 | 119,012 (37.9) | 2,371 (37.9) | 116,641 (37.9) |
| 2006-2015 | 59,728 (19.0) | 1,226 (19.6) | 58,502 (19.0) |
| **Caesarean delivery, n (%)** | 78,605 (25.0) | 1,668 (26.7) | 76,937 (25.0) |
| **Singleton, n (%)** | 305,954 (97.5) | 6,097 (97.4) | 299,857 (97.5) |
| **Firstborn, n (%)** | 123,695 (39.4) | 2,485 (39.7) | 121,210 (39.4) |
| **Birth weight, g, mean (SD)** | 3,367.1 (569.3) | 3,413.1 (547.5) | 3,366.2 (569.7) |
| **Gestational age at birth, days, median (p25-p75)** | 277 (269-285) | 277 (269-285) | 277 (269-285) |
| **Maternal age, years, mean (SD)** | 27.4 (6.2) | 27.9 (6.2) | 27.4 (6.2) |
| **Paternal age, years, mean (SD)** | 30.2 (7.0) | 30.6 (7.0) | 30.2 (7.0) |
| **Maternal education, n (%)** |  |  |  |
| ≤8 years | 35,354 (11.3) | 587 (9.4) | 34,767 (11.3) |
| 9-11 years | 48,597 (15.5) | 922 (14.7) | 47,675 (15.5) |
| 12 years | 73,454 (23.4) | 1,473 (23.5) | 71,981 (23.4) |
| 13-15 years | 51,505 (16.4) | 1,077 (17.2) | 50,428 (16.4) |
| ≥16 years | 52,562 (16.7) | 1,130 (18.1) | 51,432 (16.7) |
| **Census block group <150% poverty, n (%)** |  |  |  |
| 1<sup>st</sup> tertile (poorest) | 104,297 (33.2) | 2,046 (32.7) | 102,251 (33.2) |
| 2<sup>nd</sup> tertile | 104,702 (33.4) | 2,067 (33.0) | 102,635 (33.4) |
| 3<sup>rd</sup> tertile (richest) | 104,426 (33.3) | 2,136 (34.1) | 102,290 (33.3) |

p25, 25<sup>th</sup> percentile; p75, 75<sup>th</sup> percentile; SD, standard deviation

In our main analysis we observed a significant association between high ambient temperature in early pregnancy and an increased risk of childhood ALL (Figure 1). Specifically, for each week from the week preceding LMP and through gestational week 20, there was a positive and statistically significant association between ambient temperature and risk of ALL. The apex of the curve was in gestational week 8, where a 5 °C increase in mean weekly temperature was associated with an OR for ALL of 1.07 (95% confidence interval [CI] 1.04-1.11). Of note, there was no association between ambient temperature before pregnancy or in the second half of pregnancy and risk of ALL.

**Figure 1.**
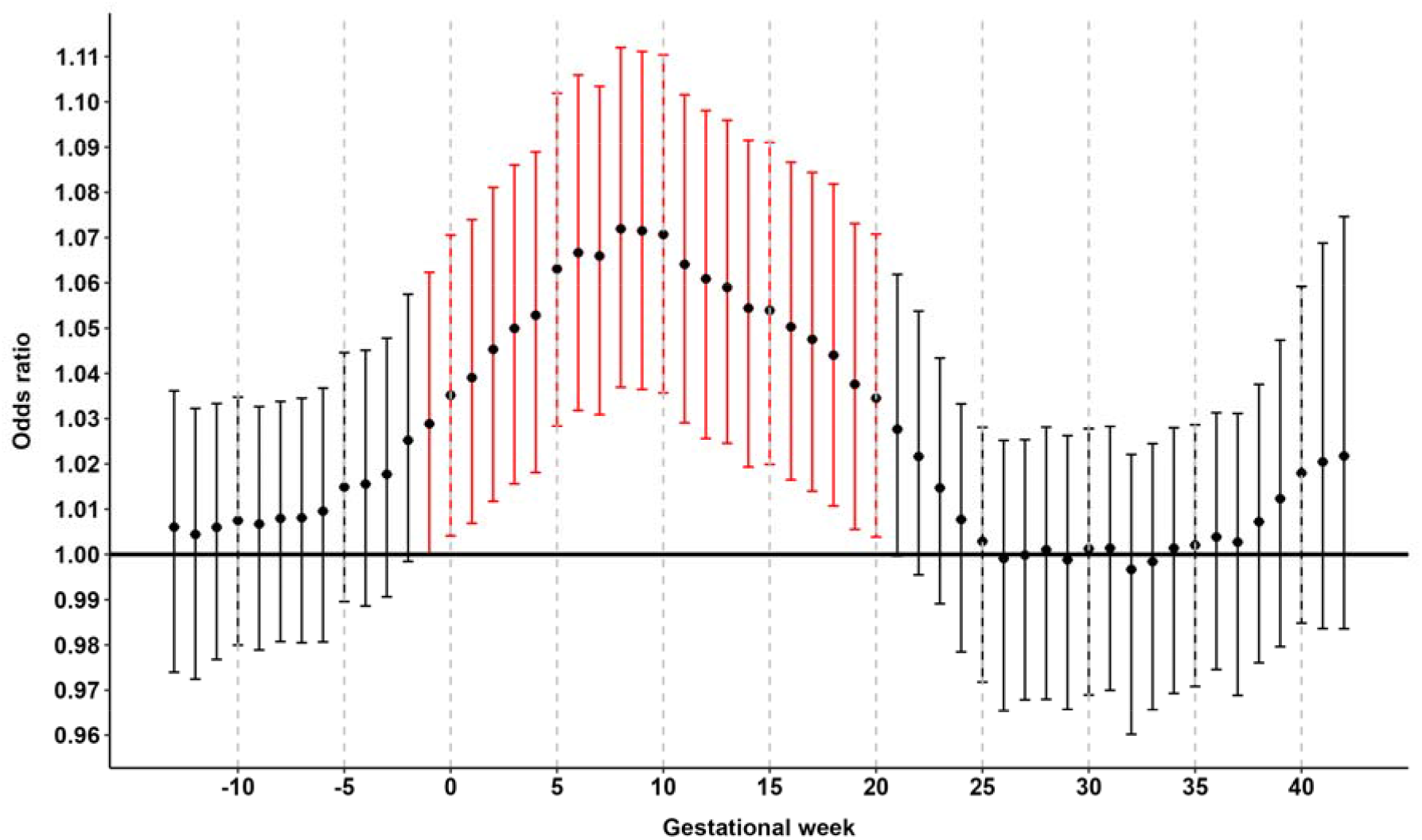
Gestational week specific associations between high ambient temperature and risk of childhood acute lymphoblastic leukemia. Results from the two-stage Bayesian meta-regression analysis of ambient temperature and risk of childhood acute lymphoblastic leukemia. Adjusted for race/ethnicity, birth order, maternal and paternal age, maternal education, neighborhood poverty, date of LMP ±7 days (i.e., seasonality and time trend), and offspring sex. Unit of exposure per 5 °C increase in mean weekly ambient temperature. Vertical bars represent 95% confidence intervals. Statistically significant associations between high ambient temperature and childhood acute lymphoblastic leukemia highlighted in red.

In subgroup analyses of gestational week 8 stratified by race/ethnicity, we observed a slightly larger effect of high ambient temperature among Latino compared with non-Latino White children (Figure 2), but the difference was not significant (p-value = 0.266).

**Figure 2.**
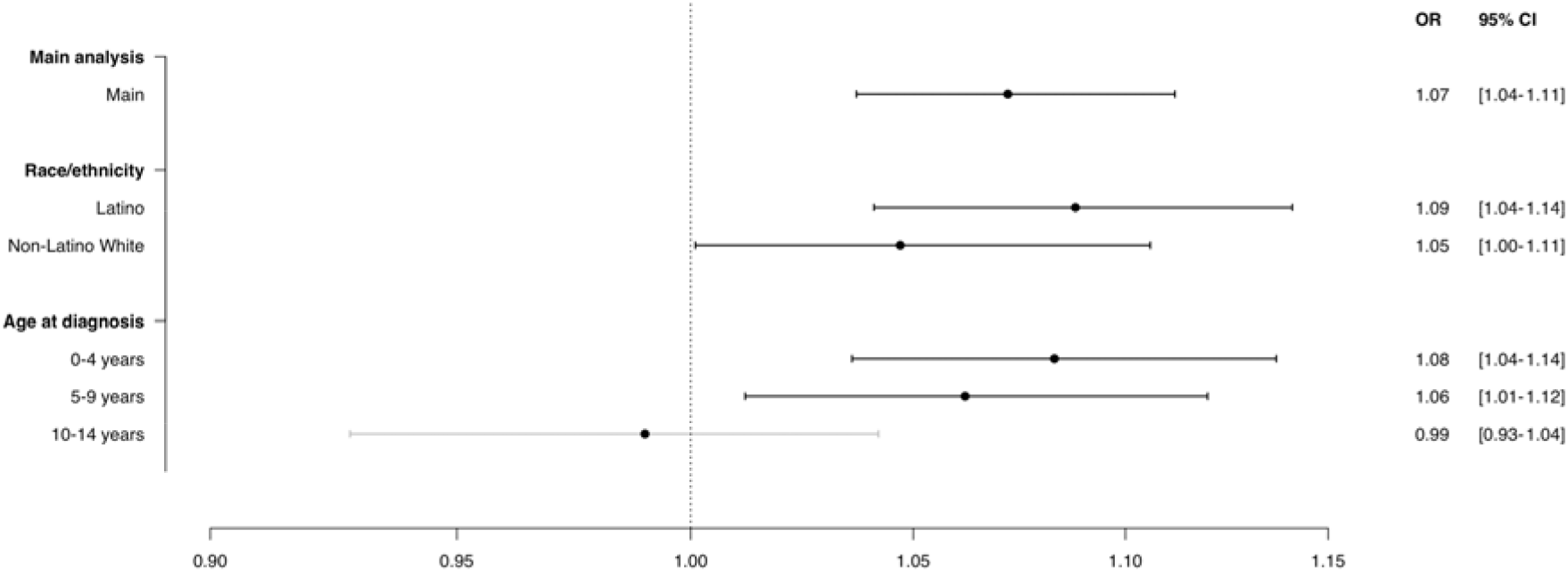
High ambient temperature in gestational week 8 and risk of childhood acute lymphoblastic leukemia, stratified by race/ethnicity and age at diagnosis. Results from the two-stage Bayesian meta-regression analysis of ambient temperature and risk of childhood acute lymphoblastic leukemia in gestational week 8 (identified as most susceptible week of exposure) and stratified by race/ethnicity and age at diagnosis. Adjusted for race/ethnicity, birth order, maternal and paternal age, maternal education, neighborhood poverty, date of LMP ±7 days (i.e., seasonality and time trend), and offspring sex. Unit of exposure per 5 °C increase in mean weekly ambient temperature. Horizontal bars represent 95% confidence intervals. Statistically significant associations between high ambient temperature and childhood acute lymphoblastic leukemia highlighted in bold horizontal bars.

Stratified by age of diagnosis, there was a comparable and pronounced association between high ambient temperature and increased risk of ALL diagnosed at 0-4 and 5-9 years of age (Figure 2; p-value of difference between the strata was 0.271). However, we observed no association between ambient temperature and risk of ALL diagnosed at the age of 10-14 years (p-values of difference between the strata 0-4 years and 10-14 years, and between 5-9 years and 10-14 years, were 0.018 and 0.074, respectively).

The non-linear analysis of ambient temperature in gestational week 8 and risk of ALL yielded a fairly linear association between the two (Figure 3). Given the wide confidence intervals, however, the analysis did not rule out a progressively increased risk per unit increase of ambient temperature, nor an attenuation of the effect of ambient temperature at very high temperatures (p-value = 0.150). Compared with a weekly mean temperature of 15 °C, mean temperatures of 20 °C, 25 °C, and 30 °C were associated with ORs for ALL of 1.23 (95% CI 1.02-1.49), 1.35 (95% CI 1.10-1.66), and 1.38 (95% CI 1.08-1.76), respectively.

**Figure 3.**
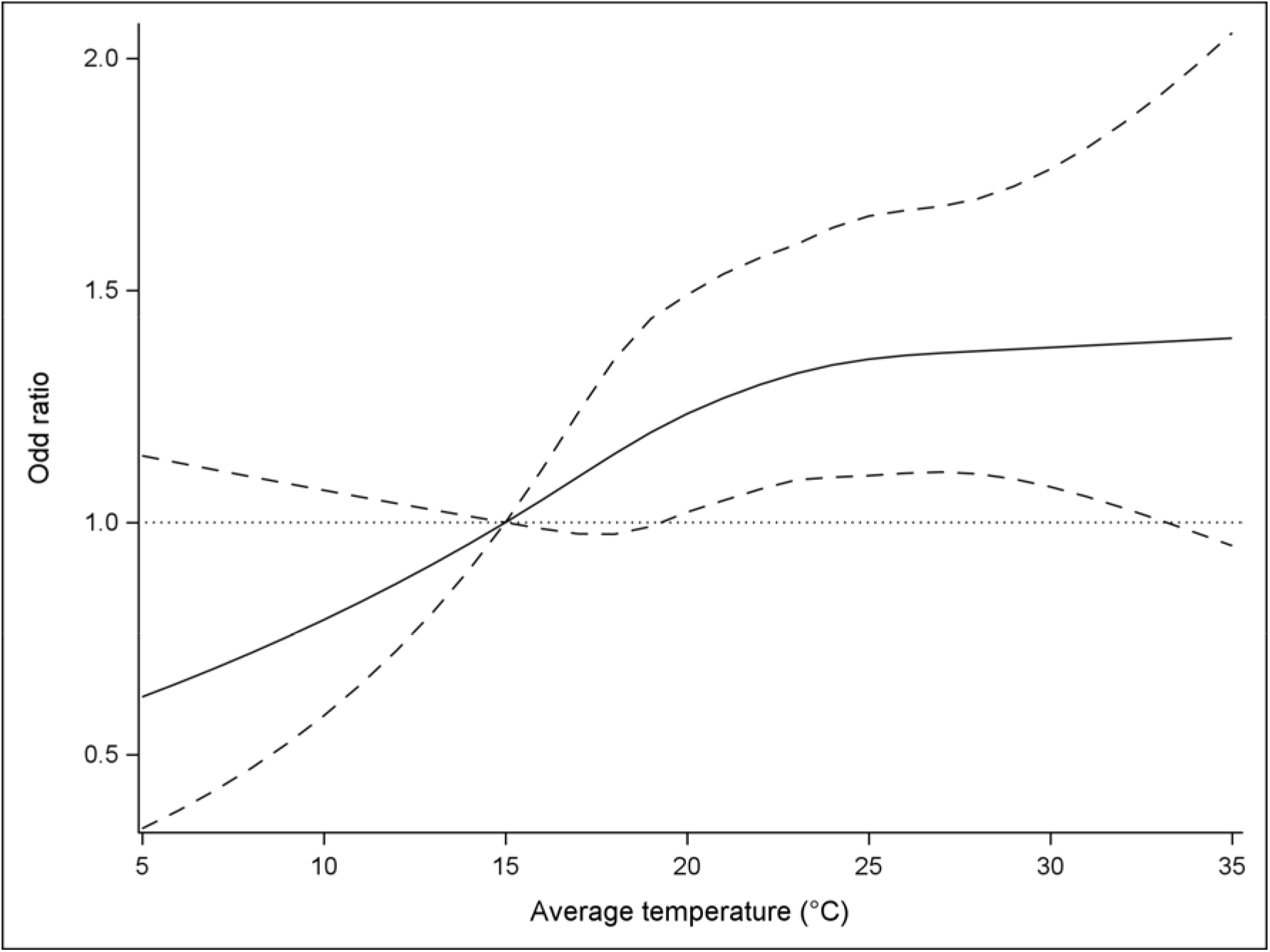
Non-linear analysis of ambient temperature and risk of childhood acute lymphoblastic leukemia in gestational week 8. Results from cubic spline analysis with 5 knots. Mean weekly ambient temperature of 15 °C used as reference. Adjusted for race/ethnicity, birth order, maternal and paternal age, maternal education, neighborhood poverty, date of LMP ±7 days (i.e., seasonality and time trend), and offspring sex. Dotted lines represent 95% confidence intervals.

Finally, we evaluated the critical windows of ambient temperature exposure using the secondary matched dataset controlling for residential address, while allowing for seasonal effects (Supplementary Figure 2). As with the main analysis, we observed a positive association between high ambient temperature in early pregnancy and risk of ALL, with a peak in gestational week 0 (OR 1.03 [95% CI1.01-1.04]).

## DISCUSSION

In this first study to evaluate the potential association between ambient temperature during pregnancy and risk of cancer in the offspring, we observed a robust association between high ambient temperature in early pregnancy and risk of childhood ALL in a set of models accounting for important confounders, exposure contrast by time and space, and evaluating a negative control exposure period. The strongest association between high ambient temperature and risk of ALL was observed in gestational week 8, in both Latino and non-Latino White subjects.

Possible biological mechanisms underlying an association between high ambient temperature in early pregnancy and the risk of childhood leukemia are unknown. In general, adverse pregnancy outcomes due to prolonged high ambient temperature are thought to be a result of maternal inflammatory and oxidative stress responses which in turn affect the developing fetus.^16^ Childhood ALL has a prenatal origin, as evidenced by detection of pre-leukemic cells in neonatal blood spots.^10^ Lymphopoiesis starts around gestational week 8.^14^ It is plausible that lymphopoiesis is most susceptible to external insults when tissue development is the most immature, as for most other tissues during embryonic and fetal development.^24^ It has been hypothesized that oxidative stress, among other factors, could be involved in the *in utero* leukemogenesis.^9^ Given that maternal heat stress can be a cause of increased oxidative stress,^15,16^ this may be a potential mechanism linking ambient temperature to ALL. Another potential pathway is that high ambient temperature may elevate the levels of air pollutants which in turn can also cause oxidative stress and inflammation.^15^ The proportion of the effect of ambient temperature in pregnancy on the risk of childhood ALL that is mediated through air pollution should be evaluated in future studies.

There are some studies of other stressors in pregnancy that are informative to the current setting. Urinary tract infections and viral infections in pregnancy, for instance, have been linked to risk of childhood leukemia.^18,25^ Children with early-onset ALL are more likely to be born during the winter and spring, which has been interpreted as a reflection of perinatal infectious exposures,^26–28^ but which also coincides with high ambient temperature around the time of conception and in the first trimester.

One of our sensitivity analyses evaluated a secondary matched dataset where we effectively compared pregnancies at the same location and same year but at different stages of pregnancy in that year, as opposed to the main analyses that compared pregnancies at the same stage of pregnancy at the same time but at different geographical locations. By design, the secondary matched dataset was vulnerable to confounding due to seasonal patterns (and therefore considered secondary to the other analyses). The apex of the curve of the sensitivity analysis was shifted towards an earlier week as compared with the main analysis. We interpret this shift as reflecting bias from seasonality. Of note, there is seasonal variation in air pollutants, and concentrations of nitrogen dioxide in California, for instance, tend to be the highest during the fall.^29^ As childhood ALL risk is associated with high levels of nitrogen dioxide in the first trimester,^13^ this would bias the sensitivity analysis to favor conceptions in the case-group to occur closer to the fall and thereby skewing the peak of high ambient temperature to occur prior to conception. A similar bias will be caused by the link between spring births and early-onset childhood ALL.^26^ Interpreted together with the main analysis, the sensitivity analysis strongly supports a positive association between ambient temperature in first trimester and risk of ALL, and underscores that residual confounding due to risk factors related to residential address is unlikely to explain the results of the main analysis.

We observed no effect of high ambient temperatures prior to pregnancy. The length of the lag-time between heat exposure and the initiation of leukemogenesis is not known, but it is reasonable to assume that the farther away from the initiation of lymphopoiesis, the weaker the association between high ambient temperature and ALL will be. The pre-pregnancy period can thus serve as an approximate negative control exposure period, and our findings offer strong support of a robust effect of ambient temperature in the first trimester on risk of ALL.

There was a tendency of a more pronounced association between high ambient temperature and risk of ALL among Latino than non-Latino White subjects. If replicated, such differences may reflect disparate exposure to heat due to occupation and residence.^1–3^ When stratified by age of onset, we observed a strong association between high ambient temperature in pregnancy and risk of early-onset childhood ALL, but no association for late-onset childhood ALL. This is in line with research that finds that early-onset childhood ALL may have a stronger link to fetal insults compared with later onset childhood ALL.^18^

Our study has notable strengths. The population-based record-linkage between the California Cancer Registry and birth records allowed us to include close to all children with ALL born in California, minimizing selection bias. The controls were a random sample representative of the birth cohort in California each year. Some controls may in theory have moved out of California and since developed childhood ALL before 14 years of age, thus introducing control misclassification. However, childhood ALL is rare, affecting 40 per million,^7^ amounting to a maximum of a dozen misclassified controls, which is unlikely to affect our results. Recall bias is not possible because that birth record data are collected prior to the diagnosis of ALL. We used highly accurate ambient temperature data within a 1-km grid, which represents a much finer level of detail than what is most often used in studies of ambient temperature in pregnancy.^4^ In addition to robust adjustments for confounding, we carried out extensive sensitivity analyses to make sure that our estimates were not due to bias, notably by examining non-sensitive windows before pregnancy and by carrying out an alternative matching strategy to allow for additional confounding adjustment. Lastly, our sample size is large and unprecedented for an epidemiologic study of childhood ALL.

There are also inherent limitations that should be highlighted. The individual-level exposure to high ambient temperature may be modified by factors such as access to adequate air conditioning and line of work; we did not have data to evaluate these factors. Also, geocoding was based on maternal residential address at the time of birth, and in situations where subjects had moved prior to birth there would be exposure misclassification. However, residential mobility has little impact when evaluating critical exposure windows in pregnancy, and there is no reason to believe that this exposure misclassification would be differential by case/control-status, so any bias would be towards the null.^30^ In the present study, we did not have the opportunity to address possible mechanisms of action that are at play in the link between ambient temperature and ALL (e.g., epigenetic modifications); this will be an important topic for future research.

Due to climate change, high ambient temperature is expected to be more common and intense over the coming decades, in the US and worldwide.^1^ We have for the first time established a robust association between high ambient temperature during pregnancy and risk of childhood ALL. Our study is adding to a growing body of literature that underscores that high ambient temperature not only has immediate health effects, but may be a cause of future chronic diseases.

## Data Availability

Data can be obtained through application to California Department of Public Health.

## REFERENCES

1 Romanello M, Di Napoli C, Drummond P, Green C, Kennard H, Lampard P, et al. The 2022 report of the Lancet Countdown on health and climate change: health at the mercy of fossil fuels. Lancet 2022;6736:. https://doi.org/10.1016/S0140-6736(22)01540-9.

2 Castillo F, Mora AM, Kayser GL, Vanos J, Hyland C, Yang AR, et al. Environmental Health Threats to Latino Migrant Farmworkers. Annu Rev Public Health 202042:257–76. https://doi.org/10.1146/annurev-publhealth-012420-105014.

3 Jesdale BM, Morello-Frosch R, Cushing L. The racial/ ethnic distribution of heat risk-related land cover in relation to residential segregation. Environ Health Perspect 2013121:811–7. https://doi.org/10.1289/ehp.1205919.

4 Chersich MF, Pham MD, Areal A, Haghighi MM, Manyuchi A, Swift CP, et al. Associations between high temperatures in pregnancy and risk of preterm birth, low birth weight, and stillbirths: systematic review and meta-analysis. BMJ 2020371:m3811. https://doi.org/10.1136/bmj.m3811.

5 Rammah A, Whitworth KW, Han I, Chan W, Hess JW, Symanski E. Temperature, placental abruption and stillbirth. Environ Int 2019131:105067. https://doi.org/10.1016/j.envint.2019.105067.

6 Barrington-Trimis JL, Cockburn M, Metayer C, Gauderman WJ, Wiemels J, McKean-Cowdin R. Trends in childhood leukemia incidence over two decades from 1992 to 2013. Int J Cancer 2017140:1000–8. https://doi.org/10.1002/ijc.30487.

7 Feng Q, De Smith AJ, Vergara-Lluri M, Muskens IS, McKean-Cowdin R, Kogan S, et al. Trends in Acute Lymphoblastic Leukemia Incidence in the United States by Race/Ethnicity from 2000 to 2016. Am J Epidemiol 2021190:519–27. https://doi.org/10.1093/aje/kwaa215.

8 Jeon S, de Smith AJ, Li S, Chen M, Chan TF, Muskens IS, et al. Genome-wide trans-ethnic meta-analysis identifies novel susceptibility loci for childhood acute lymphoblastic leukemia. Leukemia 202236:865–8. https://doi.org/10.1038/s41375-021-01465-1.

9 Greaves M. A causal mechanism for childhood acute lymphoblastic leukaemia. Nat Rev Cancer 201818:471–84. https://doi.org/10.1038/s41568-018-0015-6.

10 Marshall GM, Carter DR, Cheung BB, Liu T, Mateos MK, Meyerowitz JG, et al. The prenatal origins of cancer. Nat Rev Cancer 201414:277–89. https://doi.org/10.1038/nrc3679.

11 Francis SS, Wallace AD, Wendt GA, Li L, Liu F, Riley LW, et al. In utero cytomegalovirus infection and development of childhood acute lymphoblastic leukemia. Blood 2017129:1680–4. https://doi.org/10.1182/blood-2016-07-723148.

12 Zhong C, Wang R, Morimoto LM, Longcore T, Franklin M, Rogne T, et al. Outdoor artificial light at night, air pollution, and risk of childhood acute lymphoblastic leukemia in the California Linkage Study of Early-Onset Cancers. Sci Rep 202313:583. https://doi.org/10.1038/s41598-022-23682-z.

13 Lavigne É, Bélair MA, Do MT, Stieb DM, Hystad P, van Donkelaar A, et al. Maternal exposure to ambient air pollution and risk of early childhood cancers: A population-based study in Ontario, Canada. Environ Int 2017100:139–47. https://doi.org/10.1016/j.envint.2017.01.004.

14 Jackson TR, Ling RE, Roy A. The Origin of B-cells: Human Fetal B Cell Development and Implications for the Pathogenesis of Childhood Acute Lymphoblastic Leukemia. Front Immunol 202112:1–10. https://doi.org/10.3389/fimmu.2021.637975.

15 Ebi KL, Capon A, Berry P, Broderick C, de Dear R, Havenith G, et al. Hot weather and heat extremes: health risks. Lancet 2021398:698–708. https://doi.org/10.1016/S0140-6736(21)01208-3.

16 Ha S. The Changing Climate and Pregnancy Health. Curr Environ Heal Reports 20229:263–75. https://doi.org/10.1007/s40572-022-00345-9.

17 Li J, Vestergaard M, Obel C, Cnattingus S, Gissler M, Ahrensberg J, et al. Antenatal maternal bereavement and childhood cancer in the offspring: A population-based cohort study in 6 million children. Br J Cancer 2012107:544–8. https://doi.org/10.1038/bjc.2012.288.

18 He J, Yu Y, Fang F, Gissler M, Magnus P, László KD, et al. Evaluation of Maternal Infection During Pregnancy and Childhood Leukemia Among Offspring in Denmark. JAMA Netw Open 20236:e230133. https://doi.org/10.1001/jamanetworkopen.2023.0133.

19 Francis SS, Wang R, Enders C, Prado I, Wiemels JL, Ma X, et al. Socioeconomic status and childhood central nervous system tumors in California. Cancer Causes Control 202132:27–39. https://doi.org/10.1007/s10552-020-01348-3.

20 Thornton MM, Shrestha R, Wei Y, Thornton PE, Kao S, Wilson BE. Daymet: Daily Surface Weather Data on a 1-km Grid for North America, Version 4 2022.https://doi.org/https://doi.org/10.3334/ORNLDAAC/1840.

21 Lipsitch M, Tchetgen Tchetgen E, Cohen T. Negative Controls: A tool for detecting confounding and bias in observational studies. Epidemiology 201021:383–8. https://doi.org/10.1097/EDE.0b013e3181d61eeb.

22 Warren JL, Kong W, Luben TJ, Chang HH. Critical window variable selection: Estimating the impact of air pollution on very preterm birth. Biostatistics 202021:790–806. https://doi.org/10.1093/biostatistics/kxz006.

23 Altman DG, Bland JM. Education and debate: Statistics Notes: Interaction revisited_LJ_: the difference between two estimates. BMJ 2003326:219.

24 Heindel JJ, Vandenberg LN. Developmental origins of health and disease. Curr Opin Pediatr 201527:248–53. https://doi.org/10.1097/MOP.0000000000000191.

25 He JR, Ramakrishnan R, Hirst JE, Bonaventure A, Francis SS, Paltiel O, et al. Maternal Infection in Pregnancy and Childhood Leukemia: A Systematic Review and Meta-analysis. J Pediatr 2020217:98–109.e8. https://doi.org/10.1016/j.jpeds.2019.10.046.

26 Sørensen HT, Pedersen L, Olsen J, Rothman K. Seasonal variation in month of birth and diagnosis of early childhood acute lymphoblastic leukemia. JAMA 2001285:168–9. https://doi.org/10.1016/j.mcna.2017.03.008.

27 András Nyari T, Kajtár P, Parker L. Seasonality of birth and acute lymphoblastic leukemia. J Perinat Med 200634:507–8. https://doi.org/10.1515/JPM.2006.101.

28 Basta NO, James PW, Craft AW, McNally RJQ. Season of birth and diagnosis for childhood cancer in Northern England, 1968-2005. Paediatr Perinat Epidemiol 201024:309–18. https://doi.org/10.1111/j.1365-3016.2010.01112.x.

29 US EPA AirData. Outdoor Air Quality Data. n.d. URL: https://www.epa.gov/air-data (Accessed February 23, 2023).

30 Warren JL, Son JY, Pereira G, Leaderer BP, Bell ML. Investigating the Impact of Maternal Residential Mobility on Identifying Critical Windows of Susceptibility to Ambient Air Pollution during Pregnancy. Am J Epidemiol 2018187:992–1000. https://doi.org/10.1093/aje/kwx335.

