## Supplementary Material for "High Ambient Temperature in Pregnancy and Risk of Childhood Acute Lymphoblastic Leukemia"

**SUPPLEMENTARY METHODS**

***Identification of Critical Windows of Exposure***

This second stage modeling resulted in estimates (i.e., posterior means), 95% credible intervals (i.e., equal tailed, quantile based), and relative importance estimates (i.e., marginal posterior inclusion probabilities) for each of the true associations of interest. CWVS was previously shown to provide improved estimation and statistical inference for identifying critical windows of susceptibility.

To ensure that the new methodology performed as expected, we carried out an additional sensitivity analysis where in each weekly analysis we randomly shuffled the observed mean temperature values across the cases and controls (i.e., effectively breaking any association between temperature and ALL risk). Under this randomization of exposure, we expected to not observe any statistically significant association from the second stage meta-regression (i.e., credible intervals that excluded one). This is also what we observed.

**SUPPLEMENTARY FIGURES**

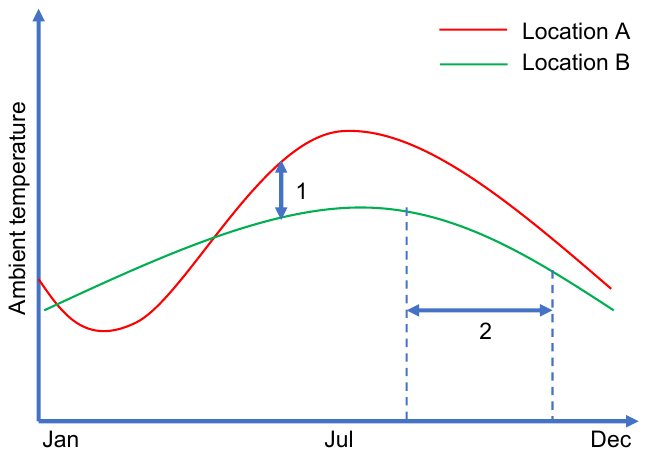

**Supplementary Figure 1.** Comparison of main and secondary matching strategies.

*Legend:* In the main matching strategy, **1**, we compared pregnancies occurring at the same time but in different locations, while in the secondary matching strategy, **2**, we compared pregnancies occurring in the same location and in the same year, but at different times during that year.

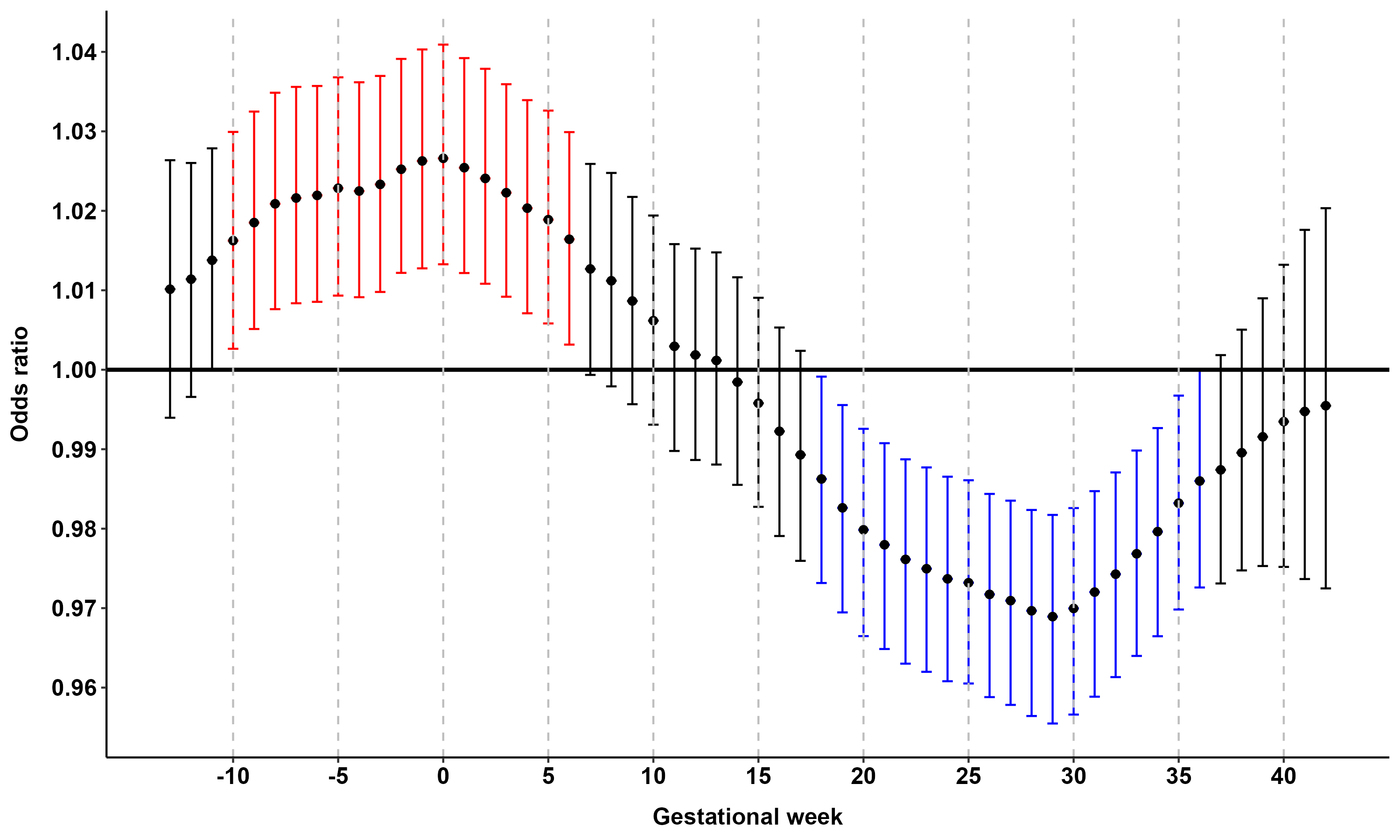

**Supplementary Figure 2.** Sensitivity analysis of gestational week specific associations between ambient temperature and risk of childhood acute lymphoblastic leukemia using the alternative dataset matched on residential address at birth.

*Legend:* Results from the sensitivity analysis using the secondary matched dataset where cases and controls were matched based on *residential address at birth within 10 km*, sex, race/ethnicity, and *year of last menstrual period* (instead of week). This analysis compared pregnancies at the same location, but at different stages of pregnancy within the same year, as compared with the main analysis which additionally adjusted for week of last menstrual period but not for geographic location. The two-stage Bayesian meta-regression analysis of ambient temperature and risk of childhood acute lymphoblastic leukemia was applied. Adjusted for race/ethnicity, birth order, maternal and paternal age, maternal education, neighborhood poverty, date of LMP +/- 365 days (i.e., seasonality and time trend), offspring sex, and place of residence within 10 km. Unit of exposure per 5 °C increase in mean weekly ambient temperature. Vertical bars represent 95% confidence intervals. Statistically significant positive and negative associations between ambient temperature and childhood acute lymphoblastic leukemia are highlighted in red and blue, respectively.
